# Integrating Health Behavior Theories to Predict COVID-19 Vaccine Acceptance: Differences between Medical Students and Nursing Students

**DOI:** 10.1101/2021.05.18.21257416

**Authors:** Hila Rosental, Liora Shmueli

## Abstract

**Background:** This study aimed to explore behavioral-related factors predicting the intention of getting a COVID-19 vaccine among medical and nursing students using an integrative model combining the Health Belief Model (HBM) and the Theory of Planned Behavior (TPB).

**Methods:** A cross-sectional online survey was conducted among medical and nursing students aged > 18 years in their clinical years in Israel between 27 August and 28 September 2020. Hierarchical logistic regression was used to predict acceptance of a COVID-19 vaccine.

**Results:** A total number of 628 participants completed the survey. Medical students expressed higher intentions of getting vaccinated against COVID-19 than nursing students (88.1% vs. 76.2%, *p* < 0.01). The integrated model based on HBM and TPB was able to explain 66% of the variance (adjusted R^2^ = 0.66). Participants were more likely to be willing to get vaccinated if they reported higher levels of perceived susceptibility, benefits, barriers, cues to action, attitude, self-efficacy and anticipated regret. Two interaction effects revealed that male nurses had a higher intention of getting vaccinated than did female nurses and that susceptibility is a predictor of the intention of getting vaccinated only among nurses.

**Conclusions:** This study demonstrates that both models considered (i.e., HBM and TPB) are important for predicting the intention of getting a COVID-19 vaccine among medical and nursing students, and can help better guide intervention programs, based on components from both models. Our findings also highlight the importance of paying attention to a targeted group of female nurses, who expressed low vaccine acceptance.

## Introduction

In March, 2020, the World Health Organization (WHO) declared COVID-19 as a global pandemic (WHO, 2020a). Vaccination is widely considered to be the most effective, long-term solution to prevent the spread of infectious diseases in general (e.g., influenza) (WHO, 2020b) and COVID-19 in particular. At the time of conducting this study (September 2020), more than 50 candidate vaccines were in the clinical evaluation stage, and no vaccine to COVID-19 was yet available. However, it is important to note that in December 2020, the U.S. Food and Drug Administration (FDA) issued the first two emergency use authorizations (EUA) for COVID-19 vaccines. Such emergency authorizations were granted to Pfizer-BioNTech and Moderna for their vaccines designed to prevent COVID-19 (The U.S. Food and Drug Administration, 2021). Nevertheless, despite vaccine availability, a significant part of the population will still not get vaccinated, partly due to a phenomenon known as vaccine hesitancy (MacDonald, 2015).

Surprisingly, vaccine hesitancy is present even among healthcare workers (HCWs), despite the great importance of vaccinating them. In fact, the CDC recommends HCWs be among the first to get COVID-19 vaccine (Persad et al., 2020). This prioritization has several reasons. First, HCWs are at an elevated risk of being exposed to COVID-19, as compared with the general public. Indeed, almost up to 50% of COVID-19 infections occur among HCWs (Chou et al., 2020). Vaccinating HCWs can therefore help not only in preventing them from being infected, but also from further infecting others, and specifically vulnerable patients. Second, vaccinating HCWs can ensure adequate workforce and protect health care capacity. Lastly, At the policy level, HCWs play a key role in providing vaccine recommendations and counseling vaccine-hesitant patients (AACN, 2021; Maltezou et al., 2019; Nguyen et al., 2020). A few recent studies that examined the intentions of HCWs to get vaccinated once a COVID-19 vaccine becomes available found that only 40%-78% of HCWs were willing to get vaccinated, with doctors presenting higher rates than nurses (Dror et al., 2020a; Gagneux-Brunon et al., 2021; Grech et al., 2020; Kwok et al., 2021; Padureanu et al., n.d.; Unroe et al., 2020; Wang et al., 2020).

Vaccinating medical and nursing students in their clinical years is also of high importance. These students are often found on the frontline in the battle against COVID-19, providing care for patients in COVID-19 departments, taking COVID-19 tests, and providing COVID-19 vaccines to patients. In addition, they play a key role in providing vaccine recommendations and counseling vaccine-hesitant patients as future professionals. Even fewer studies focused on this group of students (Barello et al., 2020; Lucia et al., 2020; Padureanu et al., n.d.), exploring their acceptance towards the novel COVID-19 vaccine. While these studies show that this group of students expresses high intention of getting vaccinated (77%-98%), none of these studies made a clear distinction nor a comparison between medical students and nursing students.

In considering the factors associated with willingness to get vaccinated against COVID-19 among HCWs, demographic and health-related predictors, as well as predictors based on theoretical behavior models should be explored.

### Factors associated with intention of getting vaccinated among HCWs

#### Demographic and health-related predictors

Recent studies addressing predictors of intention to receive COVID-19 vaccine among HCWs revealed that significantly higher proportions of males (Dror et al., 2020a; Gagneux-Brunon et al., 2021; Grech et al., 2020; Unroe et al., 2020; Wang et al., 2020), older subjects (above 60 years of age) (Grech et al., 2020; Unroe et al., 2020) or who suffer from chronic illness (Wang et al., 2020), encounter COVID-19 patients (Wang et al., 2020) or work in COVID-19 departments and being a doctor (Dror et al., 2020a) were willing to get vaccinated.

While only few studies have investigated predictors of COVID-19 vaccine acceptance, there are numerous studies reviewing predictors associated with acceptance of influenza vaccine among HCWs. In the present study, we adopted some of these related factors in addressing the current COVID-19 pandemic. Specifically, the literature reports several additional characteristics that describe HCWs who intended to get influenza vaccine. These included, religious subjects (Corace et al., 2013), those in a steady relationship or married (Corace et al., 2013; Shahrabani et al., 2009), living with dependent children (under 21 years of age), have frequent contact with the elderly (Corace et al., 2013), or have family members with chronic illness (Corace et al., 2013). Additionally, vaccine intentions could be predicted for HCWs who had higher income (Ng et al., 2020), worked at hospitals (Black et al., 2016), especially in pulmonology departments (Maltezou et al., 2010).

#### Predictors based on theoretical behavior models

Theoretical models of health beliefs and risk perception are essential tools for understanding factors motivating and inhibiting health behavior. The Health Belief Model (HBM) (Rosenstock, 1974) and the Theory of Planned Behavior (TPB) (Ajzen, 1991) are two of the most influential theories used to predict health behaviors. While HBM and TPB have been widely used for predicting behavior related to vaccination, to the best of our knowledge, they were not used in the context of COVID-19 vaccination among HCWs. The closest studies to ours dealt with the intention of HCWs to get vaccinated against seasonal influenza.

According to HBM, the intention of getting an influenza vaccine among HCWs depends on a number of factors, including: 1) perceived severity, namely, the perception of the seriousness and consequences following catching influenza for the individual and for others (e.g., loss of work time, pain and discomfort or financial); 2) perceived susceptibility, namely, risk perception of the likelihood of infection with influenza; 3) perceived benefits, namely the potential advantages of getting vaccinated against the virus (e.g., preventing the disease); 4) perceived barriers are the perceived obstacles relevant to vaccination (these can be physical such as side effects, psychological or financial) ; 5) cues to action, namely, factors that encourage a person to get vaccinated (internal factors, such as having experienced symptoms, or external factors, such as interactions with other people, information from the media or a physician’s recommendation) and 6) general health motivation (Corace et al., 2013, 2016; Dini et al., 2018; Hopman et al., 2011; Looijmans-van den Akker et al., 2009; Ng et al., 2020; Shahrabani et al., 2009).

According to TPB, the intention of getting an influenza vaccine among HCWs depends on a number of predictors, including (Corace et al., 2016; Dini et al., 2018; Godin et al., 2010; Ng et al., 2020): 1) attitude towards vaccination; 2) subjective norms, which are the social pressures that people perceive from important others encouraging them to perform the behavior; 3)Perceived Behavioral Control (PBC), which is the degree of control a person believes he has over performing the behavior (i.e., the perceived ease or difficulty in performing the behavior) (Ajzen, 1991); and 4) anticipated regret, which is the prospective feeling of positive or negative emotions after performing or not performing the behavior (Godin et al., 2010).

It is important to emphasize that HBM and TPB were also studied in the context of COVID-19 vaccination (Wong et al., n.d., 2021), but all of these studies examined the intentions of the general public and did not focus on HCWs.

The aim of this research is twofold. First, to the best of our knowledge, this is the first study to compare medical and nursing students’ intentions and attitudes towards COVID-19 vaccination. Second, when exploring factors predicting the intention to receive COVID-19 vaccine, we use an integrative model based on the HBM and TPB approaches.

## Methods

### Study design and participants

We conducted a cross-sectional online survey among nursing and medical students in their clinical years in Israel. The survey was conducted between August 27 and September 28, 2020, before the second quarantine in Israel was announced.

Participants were recruited via opportunity sampling with a minimum overall target of 300 students per profession (nursing students, medical students). Inclusion criterion were being nursing or medical students in their clinical years, and 18 years of age and older. At the beginning of the questionnaire form (see below), the respondents were informed that their participation was voluntary, they were permitted to terminate their participation at any time and that they confirmed informed consent to participate in the research. The interviewers followed a pre-defined closed-end protocol. Participants that refused to give their consent to proceed with the questionnaire and those under the age of 18 years were excluded. The questionnaire was in Hebrew.

### Ethical considerations

This study was approved by the Ethics Committee for Non-clinical Studies of Bar Ilan-University.

### Questionnaire

The following sections describe the dependent and independent variables in the questionnaire and their operationalization in this study. The parameters comprising the study measurements used to build the conceptual model are described in Figure 1. The questionnaire consisted of the following sections: (1) HBM co-variates; (2) TPB co-variates; (3) intention to receive a future COVID-19 vaccine; (4) intention to receive influenza vaccine; (5) concerns related to the COVID-19 vaccine; (6) socio-demographic co-variates, and (7) health-related co-variates. Overall, the questionnaire consisted of 55 questions and took less than 10 minutes to complete.

**Figure 1.**
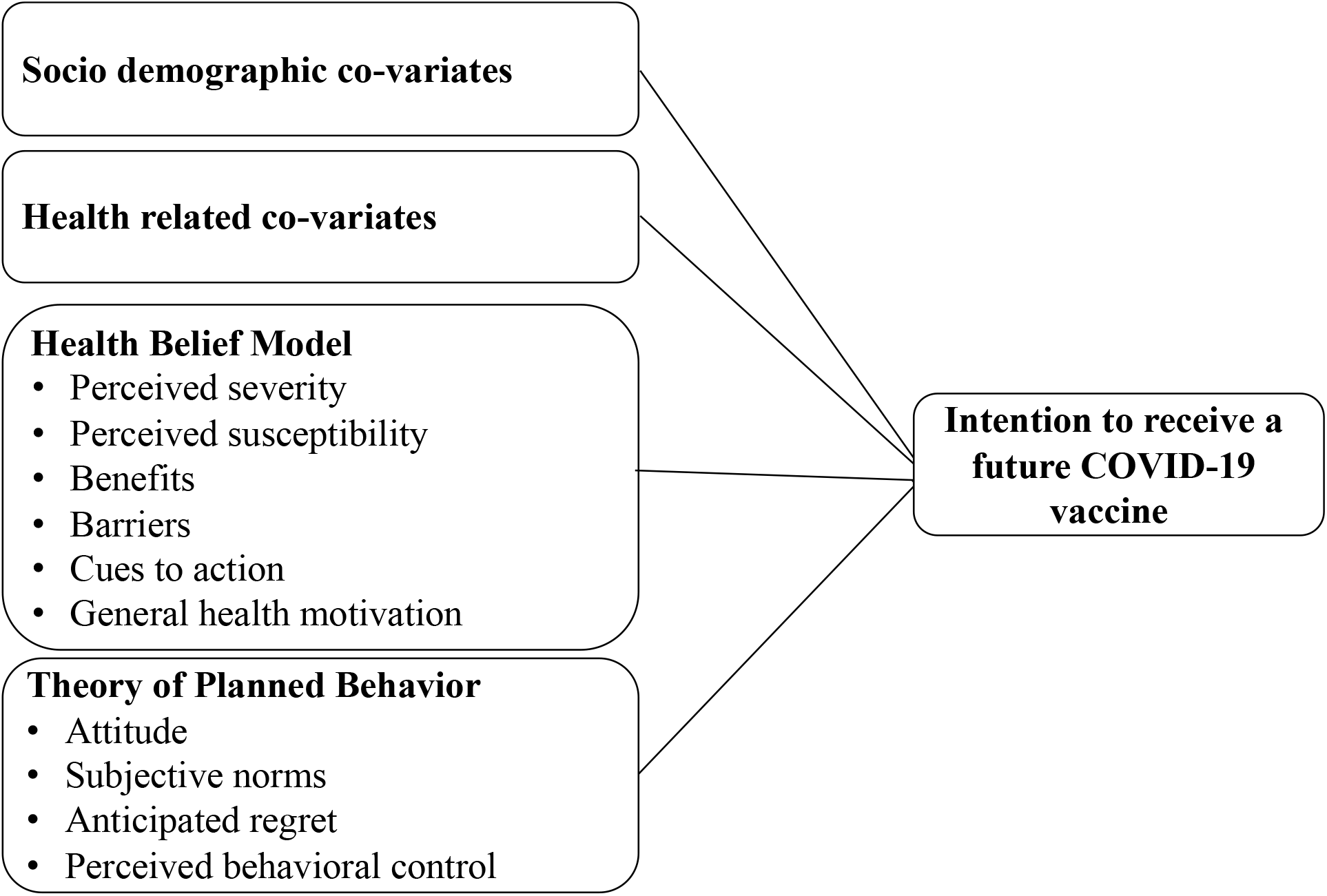
Conceptual Framework for the hypothesized predictors of intention to receive COVID-19 vaccine

### Measurement and variables

The dependent variable was the intention to receive a future COVID-19 vaccine, as measured by a one-item question on a 1-6 scale (1-strongly disagree to 6 -strongly agree). The independent variables were: (1) Socio demographic co-variates, namely, age, gender, personal status, ethnicity, and socio-economic level (based on the Israeli Central Bureau of Statistics scale and periphery level, defined by residential area); (2) health-related co-variates, such as previous influenza infection, having received influenza vaccine in the past 12 months (i.e., past behavior), suffering from a chronic disease, living with a person who belong to a high-risk group, smoking, previous or current infection with the COVID-19 virus, having contact with COVID-19 patients (at hospital, at home-care or taking samples); (3) HBM co-variates: Perceived susceptibility (included two items), perceived severity (included four items), benefits (included five items), barriers (included four items), cues to action (included five items), and general health motivation (one item); and (4) TPB co-variates: Attitude (included two items), subjective norms (included two items), PBC (included four items), and anticipated regret (included one item). Items in the HBM and TPB models were measured on a 1-6 scale (1-strongly disagree to 6 -strongly agree). Negative items were reverse scored, so that higher scores indicated higher levels of the item. Scores for each item were averaged to obtain each of the HBM and TPB independent categories.

### Statistical analyses

Data processing and analysis was done using SPSS for Windows (Version 25) software and the Process add-on for SPSS (Version 3.5) (Bolin, 2014).

A Cronbach α internal reliability method revealed that the internal consistency of HBM was Cronbach α=0.78 and that of TPB was Cronbach α =0.80. The internal consistency of the integrated model was Cronbach α=0.85. When divided into type of profession, the internal consistency of the model for nursing students was Cronbach α=0.87 and Cronbach α=0.82 for medical students.

To describe differences in variables between medical and nursing students, we conducted a series of chi-square tests (for categorical variables) and t-tests (for numeric variables).

Next, we performed a univariate analysis to identify potential predictors and meaningful interactions among the sociodemographic and health-related variables. Specifically, we performed a series of two-way analyses of variance (ANOVA) tests. For each test, a single sociodemographic or health related variable was taken, together with the profession variable, as the independent variables, and the intention of getting a COVID-19 test was taken as the dependent variable.

Finally, we performed a multivariate analysis to investigate determinants of intention to receive COVID-19 vaccine. For this purpose, we performed a hierarchical logistic regression. The intention to receive COVID-19 vaccine was used as the dependent variable measured by a one-item question on a 1-6 scale (1-strongly disagree to 6 -strongly agree). The independent variables were divided into seven blocks (A detailed description is provided in Table 3). To avoid over-complexity and possible multi-collinearity, we used the stepwise procedure from the sixth step onwards. The significance level for all analyses was set to 0.05.

## Results

### Participant characteristics

Overall, 628 respondents completed the online survey. Of these, 51% (n=321) were medical students and 49% (n=307) were nursing students. The average age of the medical students included in the sample was 28.06 years (SD=3.33), while that of nursing students was 26.04 years (SD=3.74). Among the categorical sociodemographic variables (see Table 1), significant differences were found between medical and nursing students in all cases, except for the having children variable. For example, among medical students, half of the respondents were female (n= 161) whereas among nursing students, the representation of females was significantly higher (n= 257, 83.7%). Among the health-related variables, significant differences were found for the living with someone in risk, being exposed to COVID-19 patients at work, and having received influenza vaccine in the previous season. For example, medical students reported having received influenza vaccine in the previous season at a significantly higher level, as compared to nursing students (81.6% vs. 47.6%, p<0.001). For completeness, SI Table 1 shows a comparison between medical and nursing students for each variable of the HBM and TPB models.

**Table 1.**
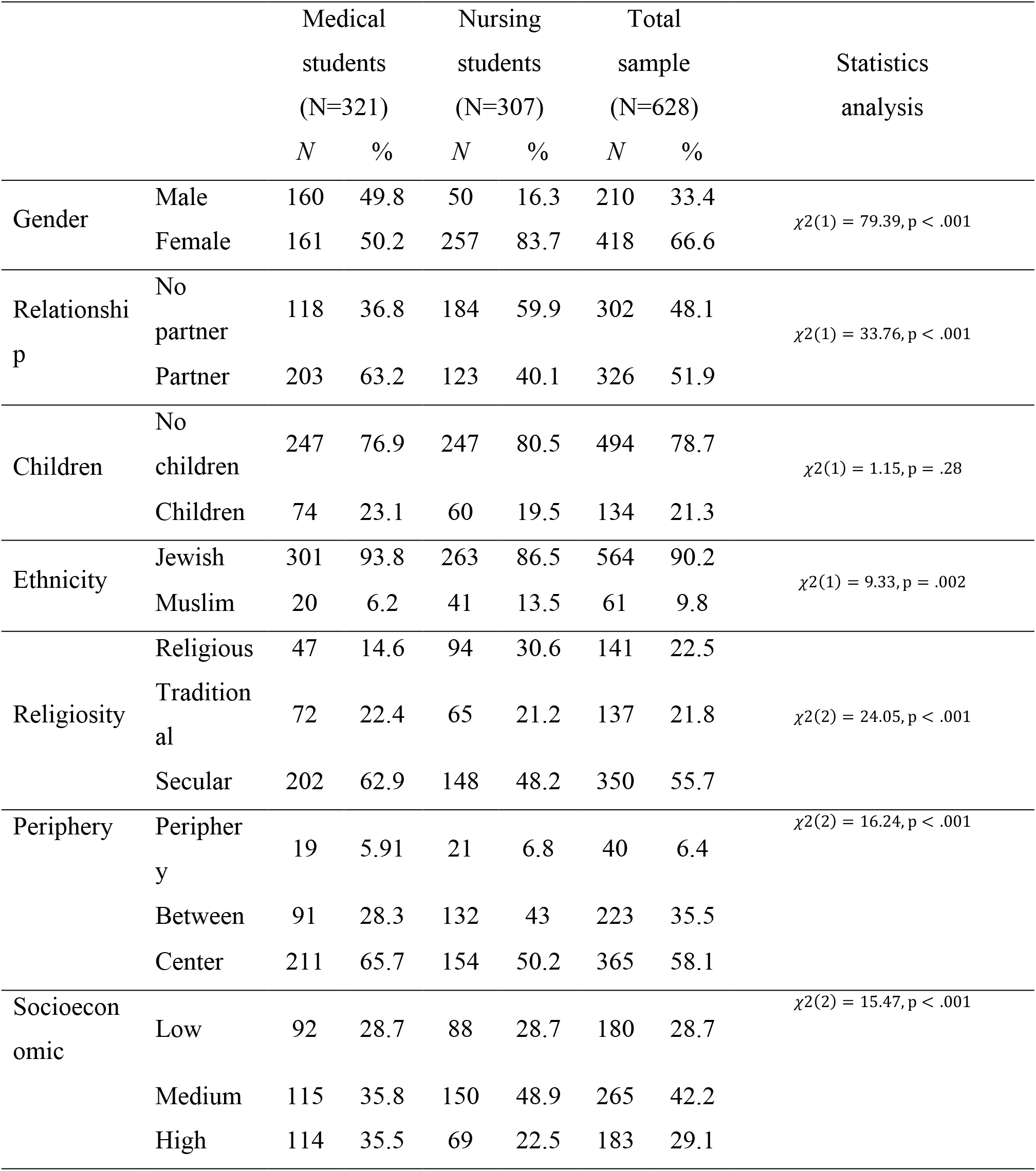
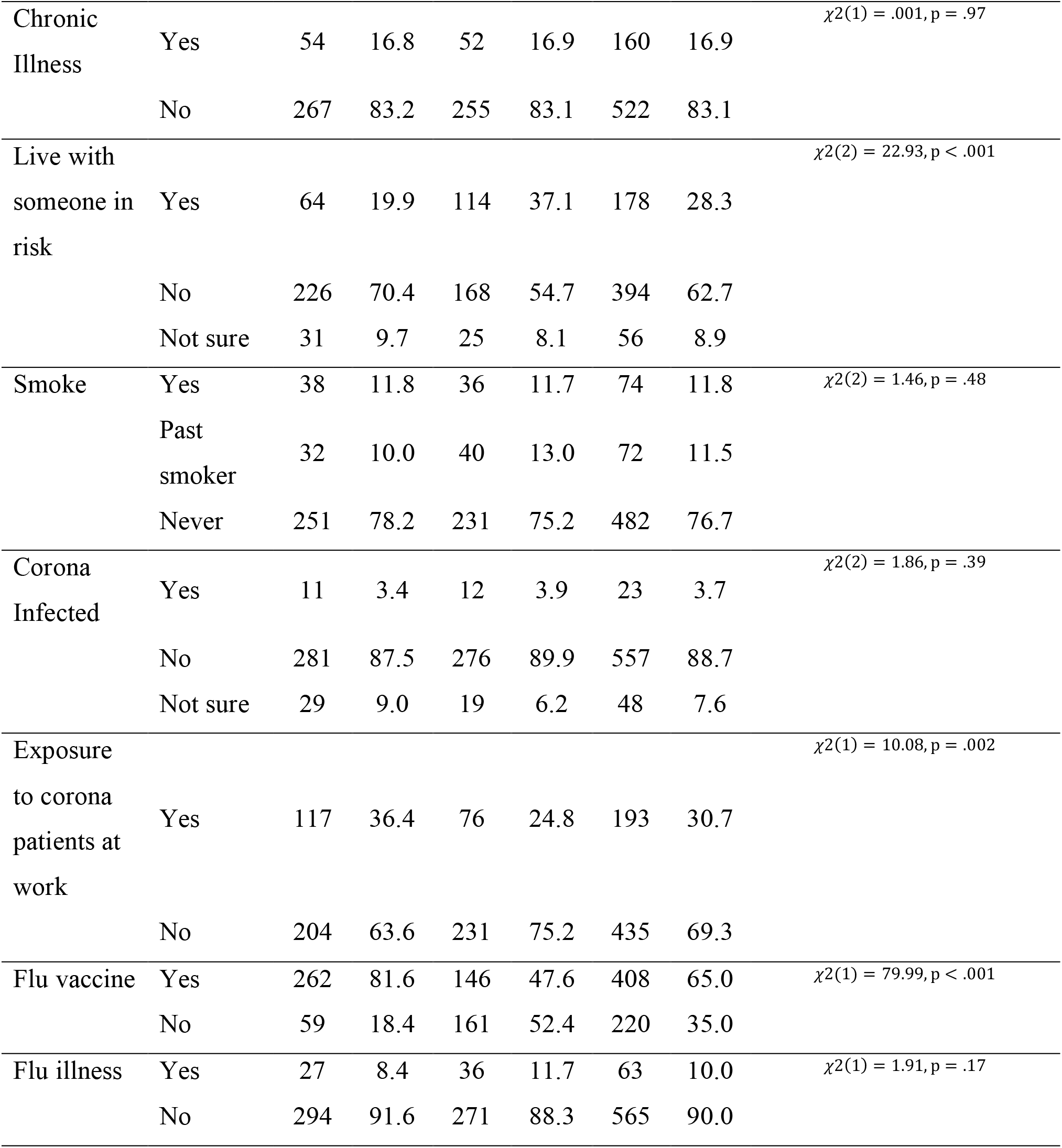
Comparison of baseline characteristics between med students and nursing students, using chi-square tests (N=628).

### Intention to receive future COVID-19 vaccine

Medical students expressed higher intentions of getting vaccinated against COVID-19 than did nursing students (88.1% vs 76.2%, p<0.01).

### Univariate analysis

Table 2 presents the significant main effects and interactions obtained when applying the two-way ANOVA tests. Recall that for each such test, the profession variable together with one of the sociodemographic or health-related variables were taken as the independent variables, and the intention of getting a COVID-19 vaccine was taken as the dependent variable. More specifically, only three main effects: socio-economic status, COVID-19 infection and previous influenza vaccination, and a single two-way interaction, profession*gender, were found significant. All other variables were not found to be significant and hence were excluded from the table. Note that although gender did not present a significant main effect, since it did present a significant two-way interaction, we kept its main effect in Table 2, as well as in the multivariate analysis that we describe below.

**Table 2.** The main significant effects and interactions obtained in the Two-Way Analyses of Variance (ANOVA) among the sociodemographic and health-related variables.

| Variable | | *m(sd)* | F | p | Partial $\eta^2$ |
| --- | --- | --- | --- | --- | --- |
| Profession | Medical students | 5.15(1.37) | 6.25 | 0.01 | .01 |
|  | Nursing students | 4.56(1.69) |  |  |  |
| Gender | Male | 5.08(1.42) | 3.04 | .08 | .01 |
|  | Female | 4.75(1.62) |  |  |  |
| Profession*Gender | Medical students. Male | 5.07(1.42) | 8.13 | .01 | .01 |
|  | Medical students. Female | 5.23(1.33) |  |  |  |
|  | Nursing students. Male | 5.12(1.47) |  |  |  |
|  | Nursing students. Female | 4.45(1.71) |  |  |  |
| SES | Low | 5.05(1.52) | 3.12 | .045 | .01 |
|  | Medium | 4.65(1.67) |  |  |  |
|  | High | 4.99(1.40) |  |  |  |
| Covid-19 infection | No | 4.88(1.54) | 4.01 | .02 | .01 |
|  | Yes | 3.96(2.03) |  |  |  |
| Previous flu vaccination | No | 4.42(1.70) | 12.10 | .001 | .02 |
|  | Yes | 5.10(1.43) |  |  |  |

### Multivariate analysis

Our integrated model included HBM and TPB variables, as well as sociodemographic and health-related variables, joined by hierarchical logistic regression so as to predict intention to receive COVID-19 vaccine. The hierarchical logistic regression coefficients and process are presented in Table 3.

**Table 3.**
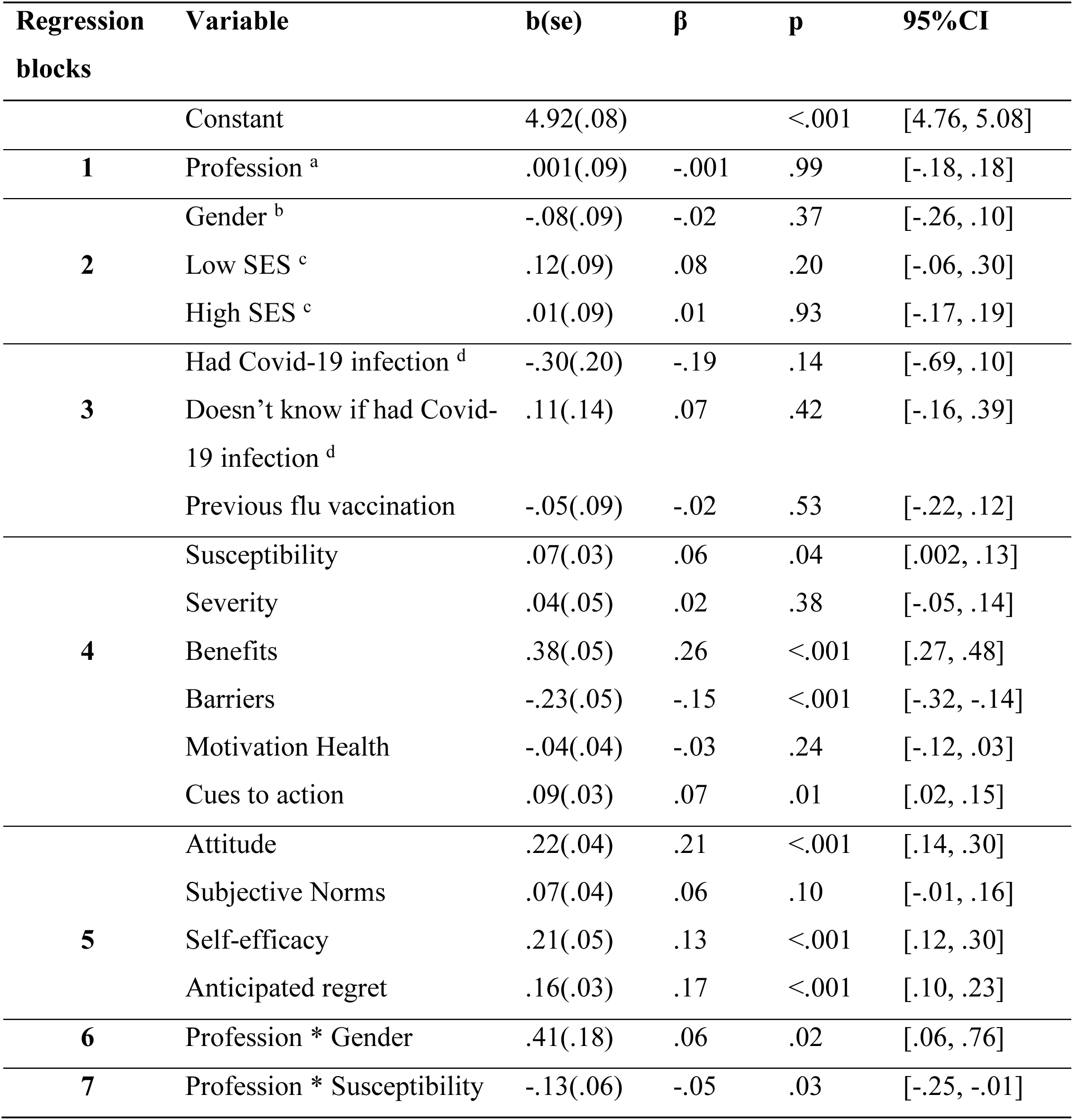
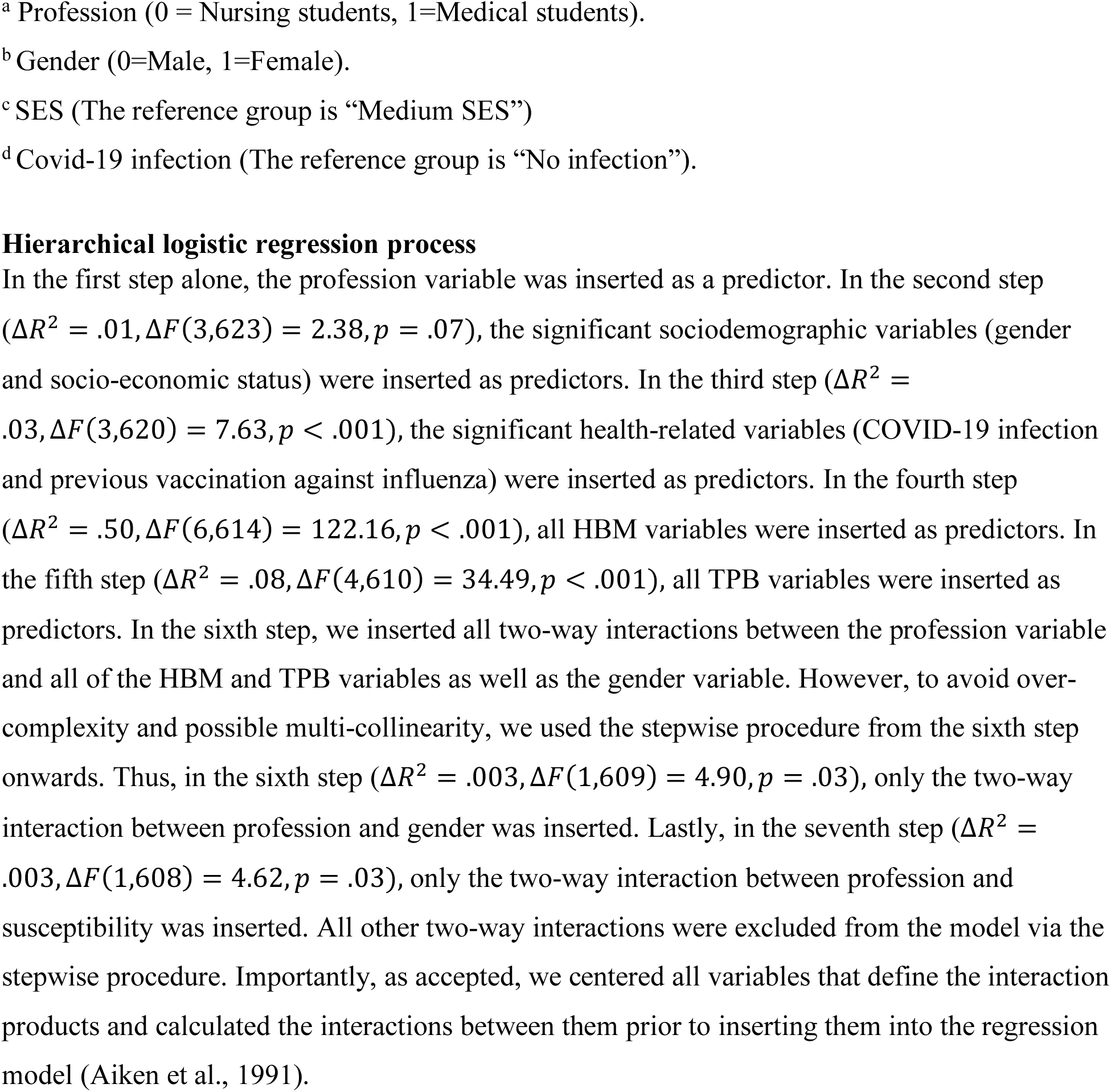
Coefficients of the final (seventh) step of the hierarchical regression. The independent variables were divided into seven blocks: 1) profession (medical or nursing student); 2) sociodemographic; 3) health-related variables that were found to be significantly correlated with intention to receive COVID-19 vaccine in the univariate analyses; 4) all HBM; 5) all TPB variables; 6) interaction between profession and gender and 7) interaction between profession and susceptibility. Here, we considered all two-way interactions between the profession variable and between HBM and TPB variables. In addition, we considered all two-way interactions that were found to be significant in the two-way ANOVA tests.

The final integrated regression model explained 66% of the variance in intention to receive COVID-19 vaccine (*R*^2^ = .66). In the first step alone, the profession variable was inserted as a predictor. In the second step, the significant sociodemographic variables (gender and socio-economic status) were inserted as predictors. In the third step, the significant health-related variables (COVID-19 infection and previous vaccination against influenza) were inserted as predictors. In the fourth step, all HBM variables were inserted as predictors. In the fifth step, all TPB variables were inserted as predictors. In the sixth step, we inserted all two-way interactions between the profession variable and all of the HBM and TPB variables as well as the gender variable. However, to avoid over-complexity and possible multi-collinearity, we used the stepwise procedure from the sixth step onwards. Thus, in the sixth step, only the two-way interaction between profession and gender was inserted. Lastly, in the seventh step, only the two-way interaction between profession and susceptibility was inserted. All other two-way interactions were excluded from the model via the stepwise procedure. Importantly, as accepted, we centered all variables that define the interaction products and calculated the interactions between them prior to inserting them into the regression model (Aiken et al., 1991).

From HBM, perceived susceptibility, benefits, and cues to action, and from TPB, attitude, self-efficacy and anticipated regret, were all positively significant predictors of intention of getting a COVID-19 vaccine. At the same time, barriers were a significant negative predictor of intention of getting a COVID-19 vaccine. Lastly, the two interaction terms (i.e. profession*gender and profession*susceptibility) were both found to also be predictors of intention of getting a COVID-19 vaccine. When broken into simple slopes, the interaction between gender and profession revealed the following: Males had a higher intention of getting vaccinated, relative to female only among nursing students (*b* = *−*.29, *se* = .15, *p* = .04, 95%*CI*[*−*.58, *−*.001]). Simple slope analysis between profession and susceptibility interaction revealed that only for nursing students, susceptibility is a positive predictor of intention of getting vaccinated (*b* = .14, *se* = .05, *p* = .003, 95%*CI*[.05, .22]).

### Concerns regarding the COVID-19 vaccine

The main concerns regarding the COVID-19 vaccine of the respondents are provided in Table 4. Nursing students had significantly higher concerns than did medical students (all p values <.01). Specifically, nursing students had significantly higher preference for natural immunity than did medical students (M=3.63, SD=1.64 vs. M=2.34, SD=1.42, p<0.1). The most common concern was the safety and quality of the vaccine (M=4.52, SD=1.49).

**Table 4.** COVID-19 vaccine concerns among medical and nursing students (N=628)

|  | Medical students<br>(N=321) |  | Nursing<br>students<br>(N=307) |  | Total sample<br>(N=628) |  | t value |
| --- | --- | --- | --- | --- | --- | --- | --- |
|  | *M* | *SD* | *M* | *SD* | *M* | *SD* |  |
| Temporary solution | 3.79 | 1.29 | 4.22 | 1.22 | 4.00 | 1.27 | 4.27** |
| Safety and quality | 4.36 | 1.54 | 4.68 | 1.42 | 4.52 | 1.49 | 2.65** |
| Not tried on others | 4.17 | 1.59 | 4.64 | 1.43 | 4.40 | 1.53 | 3.88** |
| Low efficiency | 3.93 | 1.42 | 4.29 | 1.40 | 4.11 | 1.43 | 3.12** |
| Natural Immunity is<br>preferable | 2.34 | 1.42 | 3.63 | 1.64 | 2.97 | 1.65 | 10.54** |

\*\*p<.01.

## Discussion

The present study examined acceptance rates and predictors of medical and nursing students intention to receive a future COVID-19 vaccine. HCWs, including students, play an important role in the efforts against COVID-19, and better understanding of their attitudes towards COVID-19 vaccination is of high importance. To the best of our knowledge, this is the first study to compare medical and nursing students’ intentions and attitudes towards COVID-19 vaccination, and to use an integrative model combining the HBM and TPB approaches.

The overall intention to receive COVID-19 vaccine found in the present study was very high. However, medical students expressed higher intention of getting vaccinated against COVID-19 than did nursing students (88.1% vs 76.2%). These results are compatible with the findings of Dror et al., who showed that with regard to professionals (i.e., not students), vaccine acceptance among doctors (78%) was significantly higher than among nurses (61%) (Dror et al., 2020a).

We examined several predictors for the intention to receive COVID-19 vaccine, which were not previously studied in the context of COVID-19 vaccine acceptance among medical and nursing students, such as: socioeconomic status, periphery region. We also examined the interaction between profession and gender and found that males had higher intentions of getting vaccinated than females, but only among nursing students. This finding is in line with previous studies in the context of seasonal influenza, who showed that males had higher intentions to get vaccinated than females among nurses (Flanagan et al., 2020; Zhang et al., 2012). Our findings highlight the importance of focusing on female nurses when developing a targeted approach aimed at increasing vaccination rates, especially considering the fact that in our study, female nurses constitute more than 80% of that profession.

We also considered the use of risk perception models by considering an integrated approach involving both HBM and TPB, in addition to socio-demographic and health-related variables. The resulting model was able to explain 66% of the variance in the intention to receive COVID-19 vaccine among medical and nursing students, which was considerably higher than models based solely on HBM or TPB. This finding is consistent with previous studies conducted in the context of influenza, who reported that combined models can predict vaccination intentions and actual vaccination rates to a much greater extent than can each model alone (Corace et al., 2013, 2016; Ng et al., 2020).

According to HBM, susceptibility, benefits, cues to action and barriers were found to be significant predictors of the intention to receive COVID-19 vaccine. Perceived susceptibility was identified as an important factor influencing the intention of getting vaccinated against COVID-19 (Wong et al., n.d.). Moreover, we examined the interaction between profession and susceptibility, and found it to be a good predictor for the intention to receive COVID-19 vaccine among nursing students, but not among medical students. This finding highlights the need to increase the awareness of nursing students to their higher risk of being exposed to COVID-19.

Several cues to action have been presented in the literature as internal or external triggers which may signal intention of getting vaccinated. Our study shows that medical and nursing students were more motivated to get vaccinated if they were recommended to do so by their family, friends, colleagues, supervisor or GP. This finding highlights the importance of recommendations and encouragement from supervisors in healthcare organizations, as was also pointed out by previous studies in the context of influenza (Corace et al., 2013; Hopman et al., 2011).

Regarding benefits, we found that those who intend to receive the vaccine see high perceived benefits in obtaining the COVID-19 vaccine in terms of protecting themselves and others. This is similar to previous studies who showed that vaccine acceptance relies on a personal risk–benefit perception (Dror et al., 2020b), as well as a means to prevent transmission to patients and reducing the spread of the disease in general (Corace et al., 2013, 2016). Avoiding absence from work was found as another motivation of getting vaccinated. This is not surprising as the salary of medical and nursing students is typically not high, and therefore each working day is perceived as a high benefit (Hopman et al., 2011; Looijmans-van den Akker et al., 2009).

Regarding barriers, our results were similar to those of previous studies conducted among HCWs. Specifically, concerns regarding the safety aspect of vaccination and adverse side effects, expressed fear of needles and pain, or lack of time, were associated with lower intentions of getting vaccinated (Corace et al., 2013; Godin et al., 2010). When asked about concerns regarding the vaccine, nursing students had significantly higher level of concerns about safety and side effects than did medical students. Nursing students also significantly preferred natural immunity more than medical students. Previous studies mentioned that some HCWs do not want to be vaccinated in the first round and would prefer to wait and see if there are any side effects (Shekhar et al., 2021). A possible explanation for these concerns is the lack of information regarding the effectiveness, safety and quality of the COVID-19 vaccine at the time of conducting this study. Nevertheless, this finding highlights the importance of providing front-line nursing and support staff with up-to-date information on the COVID-19 vaccine, given the high degree of trust placed in them by patients. Moreover, additional educational efforts such as explanatory campaigns should be invested in nursing schools, which should include information about efficacy, safety and side effects of the vaccine.

According to our study, perceived severity and general health motivation were not found to be significant predictors. This result is consistent with previous studies reporting that severity of the disease was a much less significant predictor (Corace et al., 2016; Ng et al., 2020; Wong et al., n.d.).

According to TPB, attitude, self-efficacy and anticipated regret were found to be significant predictors of the intention to get a COVID-19 vaccine. This finding is consistent with previous studies in the context of influenza vaccination, showing that positive attitude toward the vaccine was a strong predictor of the intention to get vaccinated (Corace et al., 2016; Ng et al., 2020). Since HCWs and students serve as a role model to patients, it is important that they maintain a positive attitude towards the vaccine, as this attitude is reflected to the patients.

Finally, our study also addressed a concern with influenza vaccine at the time of the COVID-19 pandemic. In this study, past vaccinations against seasonal influenza was correlated with the decision to get vaccinated against COVID-19. This is in agreement with previous studies that examined the intention to get a COVID-19 vaccine among the general public (Sherman et al., 2020; Thunstrom et al., 2020).

### Study limitations

This study has several limitations that should be recognized when interpreting the results reported here. First, there is the time of distribution of the questionnaire. Specifically, the study was conducted before the vaccine was available. At that point, information on vaccine efficiency and safety were not definite. It is possible that were the questionnaires distributed in December 2020, the degree of reporting of intent to vaccinate would have been different as the vaccine became available. Second, this study relies on self-reported questionnaires rather than objective measurement of actual vaccination, which is subjective in manner and can lead to a bias. Additionally, although the questionnaire was anonymous, it is possible that respondents answered in a manner that would allow them to be viewed more favorably, especially due to their role in the healthcare system, and therefore a social-desirability bias might be present.

## Conclusions

We found that the intention of getting vaccinated among medical students was higher than among nursing students. Moreover, we found that among nursing students, the intention of males to get vaccinated was higher than that of females. These findings highlight the importance of paying attention and establishing intervention plans to deal with the latter group of female nurses, who expressed considerably low vaccine acceptance.

The use of HBM and TPB models is important for health policy makers and healthcare providers and can help better guide intervention programs. Specifically, we found the following predictors for the intention to receive COVID-19 vaccine: high perceived susceptibility, benefits, barriers and cues to action, attitude, self-efficacy and anticipated regret. In particular, we believe that emphasizing the benefits of vaccination, at the same time as decreasing barriers, might have a great impact on vaccination rates.

The most common concern regarding the vaccine was safety and quality. For these reasons, we presume that promoting vaccination campaigns that address the safety of the vaccine will have greater success than discussing the severity of the disease.

## Data Availability

N/A

## Abbreviations

CDC: Center for Disease Control and prevention
EUA: Emergency Use Authorization
FDA: U.S. Food and Drug Administration
HBM: Health Belief Model
HCWs: Healthcare Workers
PBC: Perceived Behavioral Control
TPB: Theory of Planned Behavior

